# National Institute for Health Research Health Informatics Collaborative (NIHR HIC): Development of a Pipeline to Collate Electronic Clinical Data for Viral Hepatitis Research

**DOI:** 10.1101/2019.12.16.19015065

**Authors:** David A Smith, Tingyan Wang, Oliver Freeman, Charles Crichton, Hizni Salih, Philippa C Matthews, Jim Davies, Kinga A Várnai, Kerrie Woods, Christopher R. Jones, Ben Glampson, Abdulrahim Mulla, Luca Mercuri, A. Torm Shaw, Lydia Drumright, Luis Romão, David Ramlakan, Finola Higgins, Alistair Weir, Eleni Nastouli, Kosh Agarwal, William Gelson, Graham S. Cooke, Eleanor Barnes

**Author notes:** Equal contributions (David A Smith, Tingyan Wang, Oliver Freeman). **Corresponding Author:** Professor Eleanor Barnes, The Peter Medawar Building for Pathogen Research, South Parks Road, Oxford, OX1 3SY.

## Abstract

**Objective:** The National Institute of Health Research (NIHR) Health Informatics Collaborative (HIC) is a programme of infrastructure development across NIHR Biomedical Research Centres (BRCs). The aim of the NIHR HIC is to improve the quality and availability of routinely-collected data for collaborative, cross-centre research. This is demonstrated through research collaborations in selected therapeutic areas, one of which is viral hepatitis.

**Design:** The collaboration in viral hepatitis identified a rich set of data points, including information on clinical assessment, antiviral treatment, laboratory test results, and health outcomes. Clinical data from different centres was standardised and combined to produce a research-ready dataset; this was used to generate insights regarding disease prevalence and treatment response.

**Results:** A comprehensive database has been developed for potential viral hepatitis research interests, with a corresponding data dictionary for researchers across the centres. An initial cohort of 960 patients with chronic hepatitis B infections and 950 patients with chronic hepatitis C infections has been collected.

**Conclusion:** For the first time, large prospective cohorts are being formed within NHS secondary care services that will allow research questions to be rapidly addressed using real world data. Interactions with industry partners will help to shape future research and will inform patient-stratified clinical practice. An emphasis on NHS-wide systems interoperability, and the increased utilisation of structured data solutions for electronic patient records, is improving access to data for research, service improvement and the reduction of clinical data gaps.

**SUMMARY:** *What is already known?:*

- Electronic Patient Records in NHS trusts contain a wealth of routinely-collected clinical data useful for translational research. However, this data is not easily accessible to the individual NHS Trust or researchers.
- There is a shortage of detailed clinical data available for patients with viral hepatitis in the UK, in particular for patients infected with the hepatitis B or E viruses.

*What does this paper add?:*

- We present a comprehensive methodology that has been proposed, implemented and validated by the NIHR HIC for the development of a new data collection and management pipeline.
- We show that routinely-collected clinical data from patients with hepatitis C, B and E infection can be collated, integrated and made available to researchers automatically from 5 large NHS trusts.
- We describe the initial data collected from 906 hepatitis B infected patients and 1404 hepatitis C infected patients.

## INTRODUCTION

The National Institute for Health Research (NIHR) Health Informatics Collaborative (HIC) [1] was established in response to a challenge by the United Kingdom’s Chief Medical Officer Dame Sally Davies to make routinely-collected clinical data available for translational research across multiple sites. The NIHR HIC is a programme of infrastructure development across the network of 23 National Health Service (NHS) Trusts, supported by their university partners through the NIHR Biomedical Research Centres (BRCs). The programme was initially made up of five NHS Trusts hosting comprehensive NIHR BRCs; Cambridge University Hospitals NHS Foundation Trust (CUH), Guy’s and St Thomas’ NHS Foundation Trust (GSTT), Imperial College Healthcare NHS Trust (ICHT), Oxford University Hospitals NHS Foundation Trust (OUH), and University College London Hospitals NHS Foundation Trust (UCLH). The aim of the NIHR HIC programme is to improve the quality and availability of routinely collected clinical data, making it available for cross-centre collaborative, translational research. This presents both opportunities and challenges:

### Opportunities

- The United Kingdom’s unified healthcare system (the NHS) generates millions of clinical datapoints each year, which can be leveraged to improve collection of clinical information, address clinical research questions and improve patient care.
- The automated collection of data from Electronic Patient Record (EPR) systems can dramatically reduce the time and cost of data collection for research and provide opportunities for collaboration with both academic and industry partners.
- Modern machine learning techniques using neural networks require large datasets to be used effectively [2], the re-use of routinely collected data can provide a cost effective way of collating these datasets.

### Challenges

- All NHS trusts are separate organizations, responsible for the protection of the data of their own patients. To enable data to be shared across these separate organizations for research, a governance framework needed to be established.
- Each NHS trust has its own EPR, and its own set of customizations, extensions, and variations in data entry practice. Alongside the primary EPR, each trust will also have an extensive collection of departmental systems, again subject to customization and variations in practice.
- Data definitions are not all standardized.
- Not all data are collected electronically at all sites.
- Large amounts of important data are stored in free text rather than discrete values.
- Clinical practice can differ between sites.
- Data can be produced and collected differently between sites. (e.g., different laboratory methods or platforms used for tests)
- Different trusts have different levels of expertise in clinical informatics.
- Projects like the NIHR HIC require sustained investment before they start to deliver tangible results.

The NIHR HIC aims to overcome these challenges and demonstrate the value of these data for research in key therapeutic areas; the first five areas considered were viral hepatitis, ovarian cancer, critical care, acute coronary syndromes, and renal transplantation. This paper focuses on the viral hepatitis theme, which is led by OUH.

Viral hepatitis is a global health problem with an estimated 1.35 million people dying from either end stage liver disease, hepatocellular carcinoma or other viral hepatitis related diseases in 2015 [3]. The majority of these deaths are as a result of Hepatitis B and C virus infection; this is greater than tuberculosis, HIV or malaria. Unlike these other infections, the n number of viral hepatitis deaths has increased since 1990 [4]. International targets arising from the United Nations “sustainable development goals” have set a challenge for the elimination of viral hepatitis as a public health threat by the year 2030 [5,6]. As part of meeting this goal, leveraging existing clinical data is a cost-effective way to answer vital research questions. The NIHR HIC Viral Hepatitis theme aims to address key research questions (Table 1), to demonstrate the utility of the NIHR HIC methodology.

**Table 1.** Overview of specific issues to be addressed in viral hepatitis theme using the National Institute of Health Research Health Informatics Collaborative viral hepatitis theme dataset.

| **Virus** | **Example research questions to be addressed using the NIHR HIC viral hepatitis theme dataset** |
| --- | --- |
| HBV and Hepatitis delta (HDV) coinfection | • Characterising demographics and laboratory parameters for cases of chronic HBV infection.<br>• Improving use of biomarkers for monitoring and treatment stratification.<br>• Identifying individuals who control/clear HBV infection.<br>• Identifying subgroups who develop complications.<br>• Estimating incidence of HDV coinfection in the UK.<br>• Identifying determinants of HDV treatment success. |
| HCV | • Establishing sustained viral response rates for HCV therapy in the UK.<br>• Identifying factors determining treatment response in difficult to treat groups of patients. |
| Hepatitis E Virus (HEV) | • Estimating incidence of HEV infection in the UK.<br>• Identifying risk factors for HEV infection the UK. |

This paper presents a comprehensive methodology that has been proposed, implemented and validated by the NIHR HIC for the development of a new data collection and management pipeline. This development is under a comprehensive governance framework that allows data to be collated across multiple centres for collaborative research on viral hepatitis. Under this governance framework, a data collaboration involves the generation of a research-ready dataset that is broad enough to support a wide range of investigations in a specific clinical area. The dataset is assembled to the same agreed standards at each centre. The data transformations needed to achieve this, starting from patient records, are documented and shared.

## METHODOLOGY

### Governance Framework

The protocol for the collection and management of the data for the viral hepatitis theme has been reviewed and approved by South Central-Oxford C Research Ethics Committee (REF Number: 15/SC/0523). In addition, a NIHR HIC data sharing framework, covering a wide range of data and research collaborations has been signed by all participating centres. This document, in conjunction with the rest of the governance framework established by the NIHR HIC, addresses common requirements and considerations regarding data sharing between centres, contractual responsibilities, confidentiality, intellectual property, and a publications policy. This general agreement will be used to underpin individual agreements for research collaborations with third party academic and industry partners. Any collaboration with industry partners requires additional agreements, with additional governance checks by participating sites. This process has been simplified by the creation of a ‘framework industry collaboration agreement’ which simplifies the addition of individual participating sites to an industry collaboration. A scientific steering committee has been established which is made up of a representative of each participating site, and reviews and approves data requests from external collaborators.

### Development of Data Model

The development of the NIHR HIC viral hepatitis data model, which outlines the structure of the dataset and the associations between the data fields, began by the clinical leads defining data fields, required to answer the initial academic questions posed by the clinical and scientific leads across centres (Table 1). The feasibility of collecting this dataset was tested using the electronic data capture software OpenClinica [7]. Each site was required to manually complete case report forms in OpenClinica for a small subset of their patient cohorts; an assessment of the completeness of these data led to a refinement of the dataset and a second version, which was used to generate the Extensible Markup Language (XML) Schema Definition (XSD) used to define the dataset. This XSD was then further refined to remove errors and to provide a more efficient data structure.

## Data Architecture for Collection and Integration of Data

Each site within the collaboration provided a data product containing the data items outlined in the agreed dataset. Whilst each site had pre-existing data warehouses and systems for collecting, storing, and using patient data, these systems were designed to be used for patient care, administrative, and financial purposes, and were not always suitable for generating the required data product. The variety of EPR systems and LIMS used across the participating sites meant that data were often stored in different formats. This meant that at each site a data architecture outlying the flow of data from EPR systems had to be developed to allow the effective flow of data from operational systems to research systems. This required the development of data warehousing or data management infrastructure at all sites.

To allow the compilation of the full dataset, each site generated a data product that was integrated in the NIHR HIC Viral Hepatitis Central Data Repository (**Figure 1**). In some cases, these data were already structured and could be transferred directly into the data repository. Where data were stored in an unstructured (free text) format, the data had to be either manually entered or extracted (**Figure 2**). The data was then anonymised by removing patient identifiers and a data product was created using the XML format. This was securely transferred to the NIHR HIC Viral Hepatitis Central Data Repository, where the data were put into a database for queries and analysis. When a dataset is requested by a research group, the request is first reviewed and approved by the scientific steering committee, and following internal governances process, an extract of the integrated dataset is provided to the research group for their analysis.

**Figure 1:**
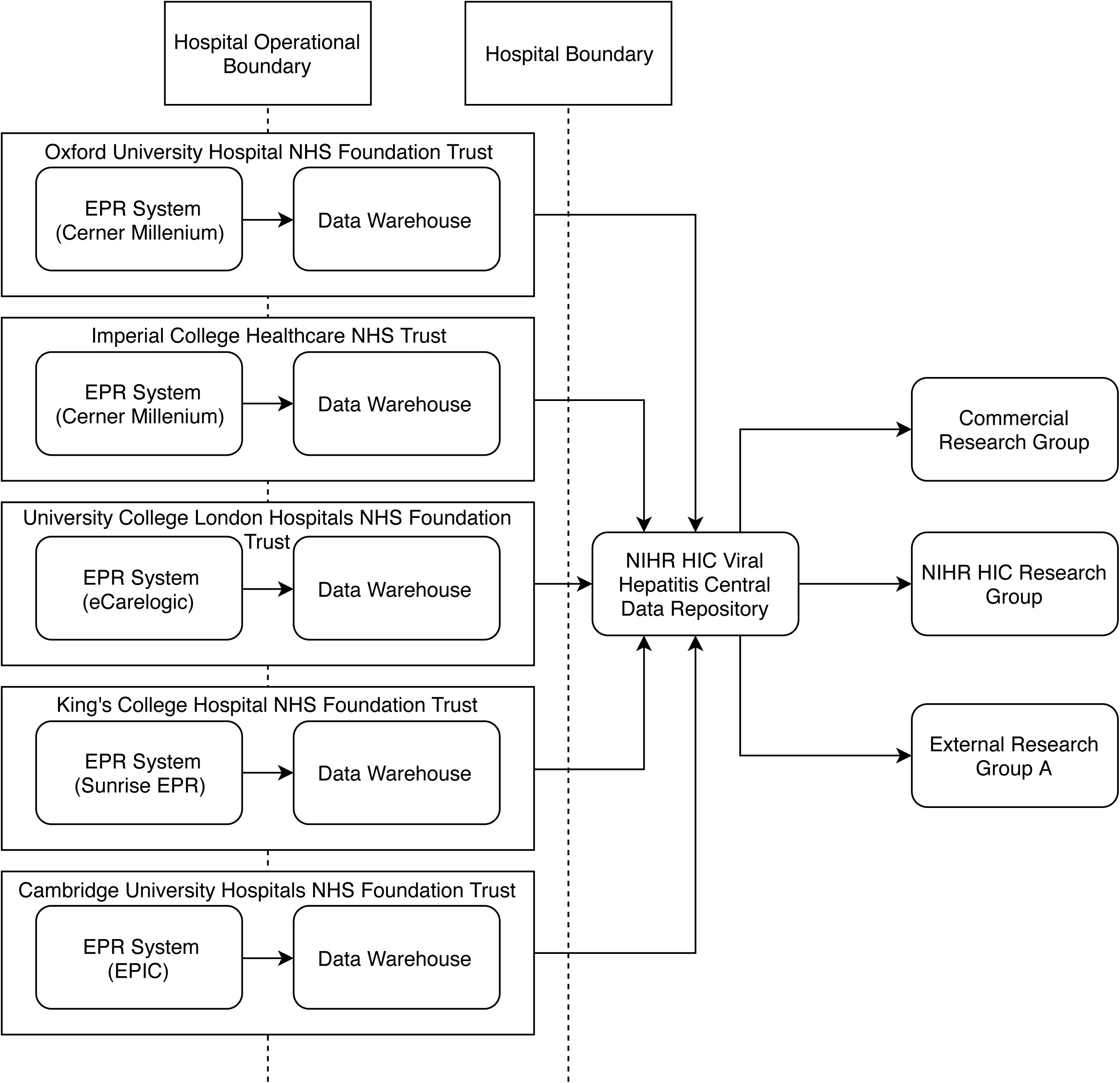
Data flow for the National Institute of Health Research Health Informatics Collaborative viral hepatitis clinical exemplar theme. For each participating site, data originating from clinical systems were used to populate a corresponding data warehouse that contains data fields in the agreed dataset for viral hepatitis research. Data in the data warehouse were transferred to the NIHR HIC Viral Hepatitis Central Data Repository. The data stored in the central data repository can be extracted and provided to internal and external research groups according to the governance process.

**Figure 2:**
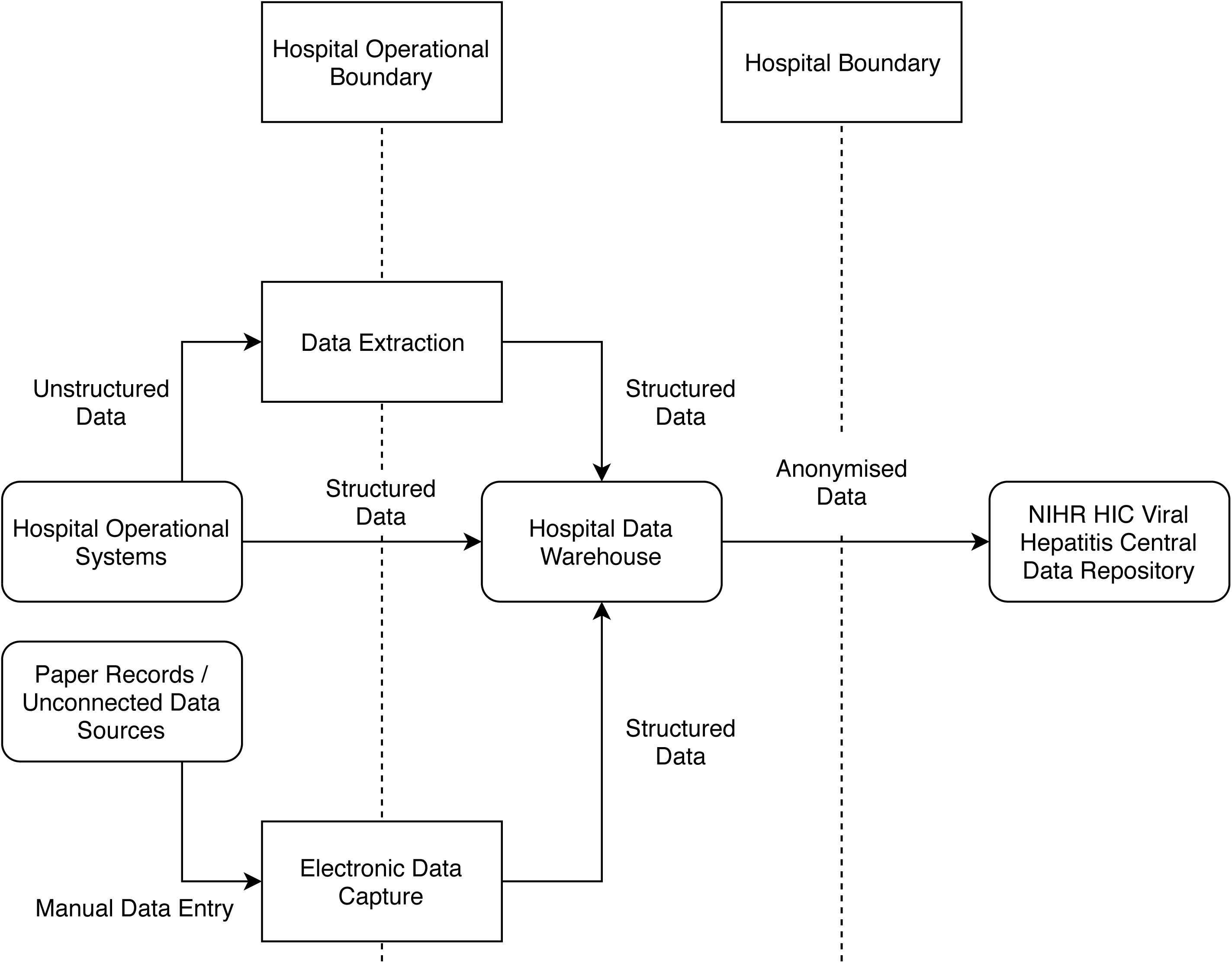
Conceptual data flow for an individual site participating in the National Institute of Health Research Health Informatics Collaborative Viral Hepatitis clinical exemplar theme. In an individual site, structured data in the hospital operational systems were directly transferred into the hospital data warehouse, and data stored in unstructured format was automatically or manually transformed to produce structured data either before or after transfer to the data warehouse. In addition, data from paper records or unconnected data sources were manually entered into a structured electronic data capture system and transferred into the data warehouse. Data was then anonymised prior to transfer to the central data repository for viral hepatitis research.

**Figure 3:**
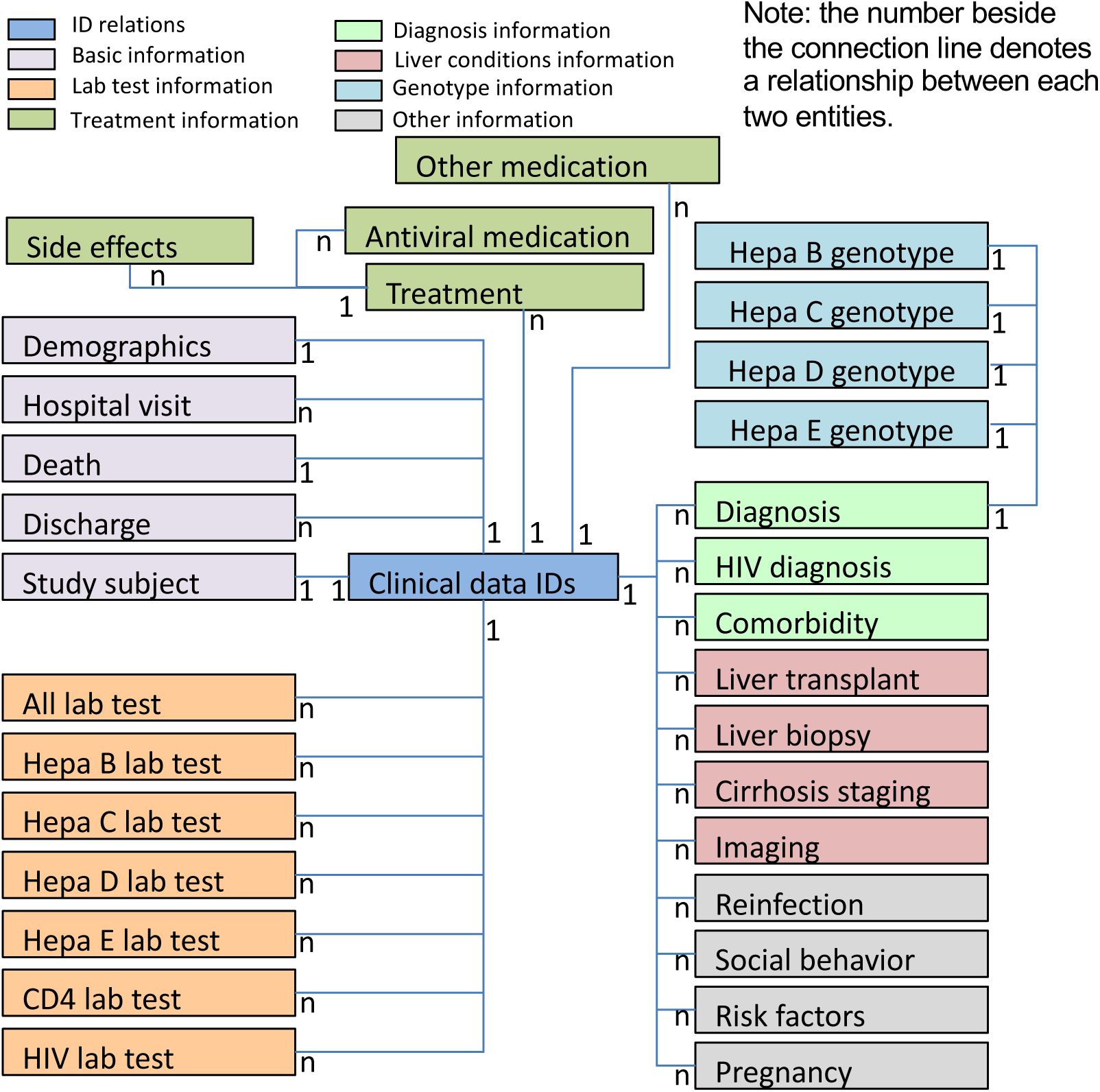
Data model overview of the National Institute of Health Research Health Informatics Collaborative Viral Hepatitis Research database. The defined dataset for viral hepatitis research includes 32 entities/tables, which can be grouped into different categories, marked in different colors. Each patient has a unique clinical data identifier that is stored in the table “Clinical data IDs”. The relationship between two entities is shown beside their connection line. For example, 1: 1 between table “Clinical data IDs” and table “Study subject” indicates that a patient can only have one study subject id; while 1: n between table “Clinical data IDs” and table “Hospital visit” indicates that a patient could possibly have multiple hospital visits.

## NIHR HIC Viral Hepatitis Central Data Repository

Anonymised data from the providing centres was transmitted to the lead centre in XML format via secure e-mail or submitted directly to the NIHR HIC Data Acquisition Management (HICDAM) system via a secure web-based front end, again in XML format. The primary service inside HICDAM is the Message Receiving, Curating, and Understanding Repository (MeRCURy), which performs the validation, processing, and storing of the submitted data.

The MeRCURy system supports two types of data validation: basic, automatic integrity checks, which must be satisfied before the data are loaded into the database, and more sophisticated, manual checks of data consistency, which are performed after the data have been loaded. The basic integrity checks involve validation against the agreed XML Schema Definition (XSD), confirming that the data are correctly formatted, together with logical checks upon the type or range of values submitted: for example, a check that any value given for the date of death is strictly later than those given for dates of treatment.

The manual checks reflect working assumptions regarding the relationships between the values of different data items submitted, for example, that a certain combination of treatments would never be used in practice. In each case, some additional information may be needed to determine whether data is incorrect, or whether the assumptions are invalid.

The system will inform data provides of the outcome of any submission. If the data submitted fail a basic integrity check, they are rejected, and a report is generated containing appropriate diagnostic information. If the data are accepted, a confirmation message is sent, and the data may then be reviewed using a secure interface provided by the LabKey application. Data can be then be explored and/or exported from the system in a variety of formats, including .xls, .xlsx, .tsv, and .csv.

## RESULTS

The NIHR HIC viral hepatitis research database has been developed and populated with data. **Error! Reference source not found**. provides an overview of the data model of the database and the relationships among different entities. The data collected falls into several categories: (a) basic information (e.g., demographics, hospital visits, death, discharge, and study sites), (b) laboratory data, (c) treatments, (d) diagnoses, (e) liver conditions, (f) hepatitis virus genotypes, and (g) other clinical information.

The database contains 32 tables for storing collected data points, and several tables for data field definitions. The final database consists of 349 data fields, split into 20 different element types. There are 203 data fields that are common to HBV, HDV, HCV and HEV whilst other data fields are specific to HBV and HDV (n=75), HCV (n=47) or HEV (n=24). The full dataset is included in supplementary data 1. To date, the database contains 3,494 patients with associated clinical information. There are 842,676 records regarding laboratory tests, 2,824 records regarding imaging data, and 8,514 records for medications (including anti-viral therapy and others), with data on comorbidities, diagnoses, genotype information, liver conditions, treatment side effects and social behavior also included.

## Chronic HBV Cohort Description

Data have been extracted from the NIHR HIC Viral Hepatitis Research database by using HBV lab test information. Patients included in the extract meet at least one of the following criteria:

a. two positive HBsAg tests, at least 6 months apart;
b. positive HBsAg and positive HBV DNA, at least 6 months apart.

The characteristics of the chronic HBV cohort of 960 patients are described in Table 2, including age, gender, race, HCV coinfection, HIV coinfection, comorbidities, the severity of liver disease, and treatment information. Among these patients, 254 patients (26.5%) have previously received medications or are currently under treatment. There are 17 patients coinfected with HDV, with data extracted based on detectable HDV viremia or anti-HDV antibody. However, there were 475 patients who had no HDV test data.

**Table 2:** Characteristics of chronic HBV cohort: NA = NOT Applicable, i.e., there are no or only a few patients who have the corresponding information; for a continuous variable, median and the interquartile range (IQR) is calculated, for a categorical variable, the number and the percentage of patients is provided. *Patients who had at least one episode of treatment recorded.

|  |  |
| --- | --- |
| **Number of patients (total)** | 960 |
| **Age at the beginning of follow-up: median (range)** | 36 (2-80) |
| **Gender (%)** |  |
| Male | 502 (52.3) |
| Female | 389 (40.5) |
| Unknown | 69 (7.2) |
| **Ethnicity (%)** |  |
| White | 106 (11.0) |
| Mixed | 11 (1.2) |
| Asian | 220 (22.9) |
| Black | 161 (16.8) |
| Other | 72 (7.5) |
| Unknown | 390 (40.6) |
| **HDV coinfection** |  |
| Yes (%) | 17 (1.8) |
| No (%) | 468 (48.7) |
| Unknown (%) | 475 (49.5) |
| **HCV coinfection** |  |
| Yes (%) | 75 (7.8) |
| No (%) | 347 (36.1) |
| Unknown (%) | 538 (56.1) |
| **HIV coinfection** |  |
| Yes (%) | 7 (0.7) |
| No (%) | 427 (44.5) |
| Unknown (%) | 526 (54.8) |
| **Documented comorbidities** |  |
| Diabetes (%) | 30 (3.1) |
| Depression (%) | 28 (2.9) |
| Chronic kidney disease (%) | 16 (1.7) |
| Coagulation disorder (%) | 22 (2.3) |
| Cryoglobulinaemia (%) | 3 (0.3) |
| Cancer (%) | 21 (2.2) |
| Liver cancer (%) | 1 (0.1) |
| Other cancer (%) | 20 (2.1) |
| **Severity of liver disease** |  |
| Cirrhosis (%) | 27 (2.8) |
| Decompensated (%) | 4 (0.4) |
| Child-Pugh score: median (IQR) | NA |
| Fibroscan stiffness: median (IQR) | 5.3 (2.4) |
| MELD score: median (IQR) | NA |
| **Patients treated (%)** | 254 (26.5) |
| **Lab tests at baseline** |  |
| ALT (IU/L) |  |
| Untreated group: median (IQR) | 29 (25) |
| Treated group: median (IQR) | 39 (37) |
| HBV DNA (log<sub>10</sub> IU/ml) |  |
| Untreated group: median (IQR) | 3.15 (1.58) |
| Treated group: median (IQR) | 4.26 (3.67) |
| HBeAg positive |  |
| Untreated group (%) | 82/706 (11.6) |
| Treated group (%) | 84/254 (33.1) |

## Chronic HCV Cohort Description

Data have been extracted for patients with a positive HCV PCR test or HCV genotype from the NIHR HIC viral hepatitis research database. The characteristics of HCV cohort of 1404 patients are described in Table 3, including age, gender, ethnicity, HCV genotype, HBV co-infection, HIV co-infection, comorbidities, the severity of liver disease, and treatment information. Among these patients, 914 patients (65.1%) had received interferon-free direct-acting antiviral (DAA) treatment by February 2018.

**Table 3:**
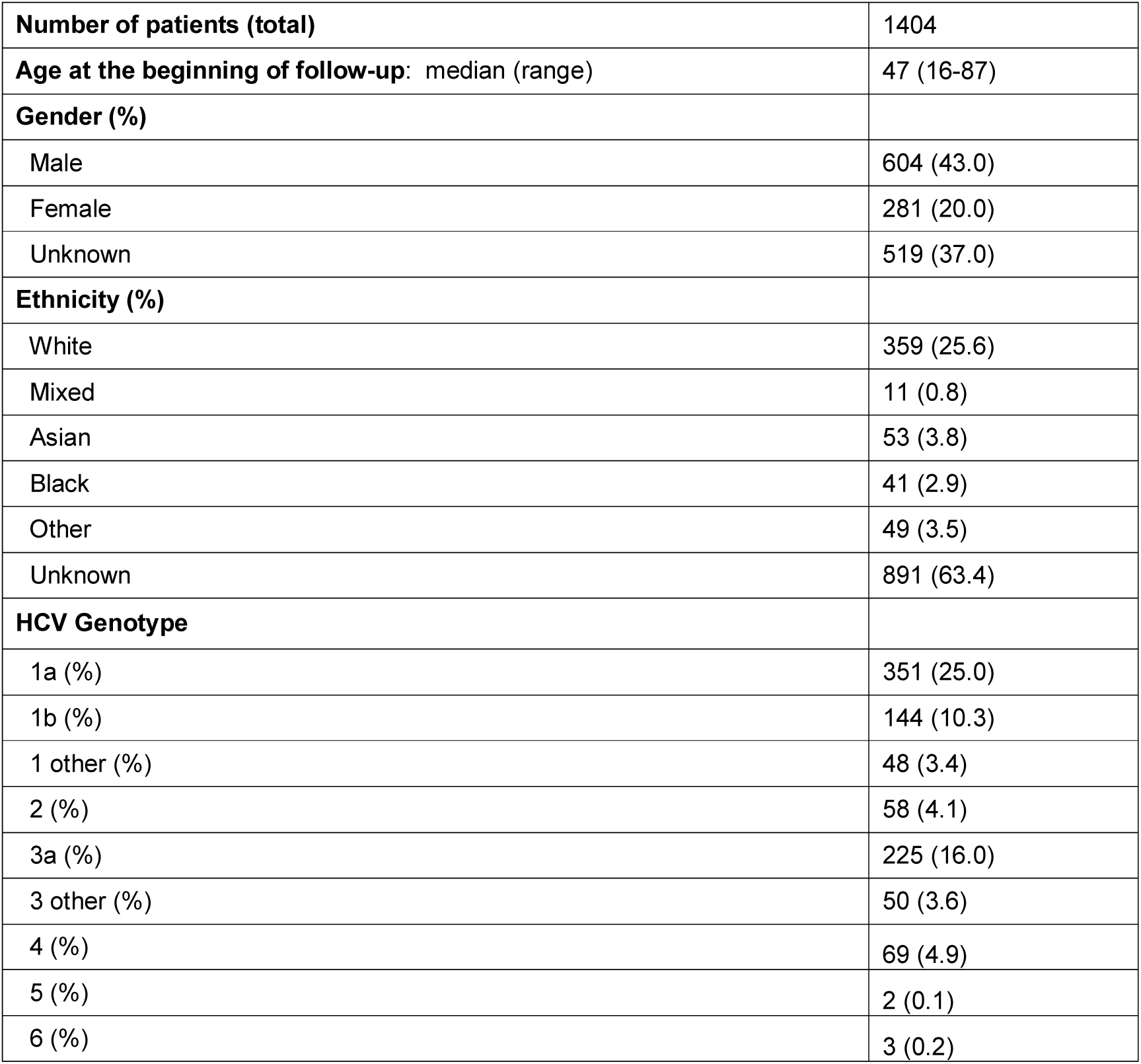
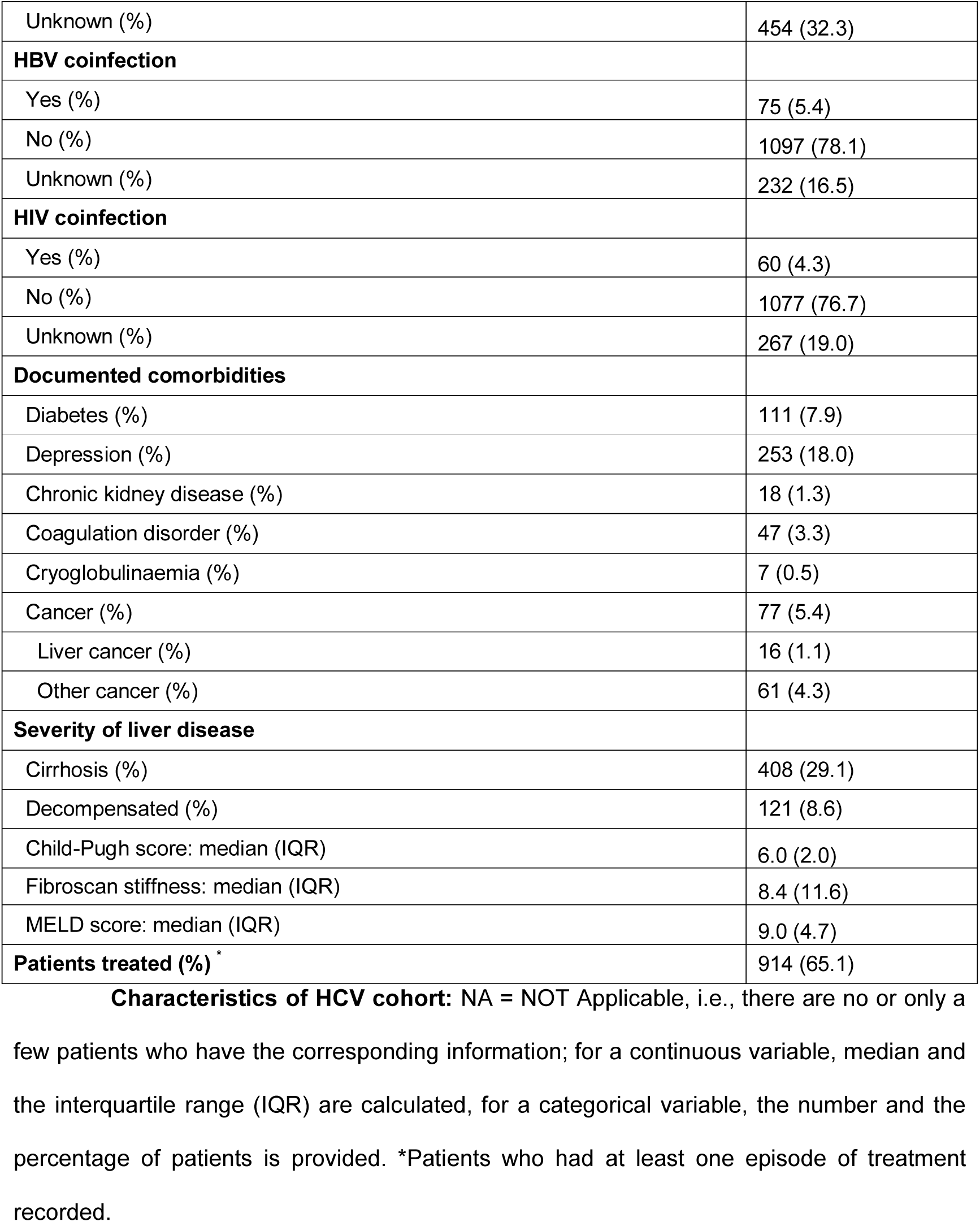
Characteristics of HCV cohort: NA = NOT Applicable, i.e., there are no or only a few patients who have the corresponding information; for a continuous variable, median and the interquartile range (IQR) are calculated, for a categorical variable, the number and the percentage of patients is provided. *Patients who had at least one episode of treatment recorded.

|  |  |
| --- | --- |
| **Number of patients (total)** | 1404 |
| **Age at the beginning of follow-up: median (range)** | 47 (16-87) |
| **Gender (%)** |  |
| Male | 604 (43.0) |
| Female | 281 (20.0) |
| Unknown | 519 (37.0) |
| **Ethnicity (%)** |  |
| White | 359 (25.6) |
| Mixed | 11 (0.8) |
| Asian | 53 (3.8) |
| Black | 41 (2.9) |
| Other | 49 (3.5) |
| Unknown | 891 (63.4) |
| **HCV Genotype** |  |
| 1a (%) | 351 (25.0) |
| 1b (%) | 144 (10.3) |
| 1 other (%) | 48 (3.4) |
| 2 (%) | 58 (4.1) |
| 3a (%) | 225 (16.0) |
| 3 other (%) | 50 (3.6) |
| 4 (%) | 69 (4.9) |
| 5 (%) | 2 (0.1) |
| 6 (%) | 3 (0.2) |
| Unknown (%) | 454 (32.3) |
| **HBV coinfection** |  |
| Yes (%) | 75 (5.4) |
| No (%) | 1097 (78.1) |
| Unknown (%) | 232 (16.5) |
| **HIV coinfection** |  |
| Yes (%) | 60 (4.3) |
| No (%) | 1077 (76.7) |
| Unknown (%) | 267 (19.0) |
| **Documented comorbidities** |  |
| Diabetes (%) | 111 (7.9) |
| Depression (%) | 253 (18.0) |
| Chronic kidney disease (%) | 18 (1.3) |
| Coagulation disorder (%) | 47 (3.3) |
| Cryoglobulinaemia (%) | 7 (0.5) |
| Cancer (%) | 77 (5.4) |
| Liver cancer (%) | 16 (1.1) |
| Other cancer (%) | 61 (4.3) |
| **Severity of liver disease** |  |
| Cirrhosis (%) | 408 (29.1) |
| Decompensated (%) | 121 (8.6) |
| Child-Pugh score: median (IQR) | 6.0 (2.0) |
| Fibroscan stiffness: median (IQR) | 8.4 (11.6) |
| MELD score: median (IQR) | 9.0 (4.7) |
| **Patients treated (%) \*** | 914 (65.1) |

## HEV Cohort Description

Data have been extracted from the NIHR HIC viral hepatitis research database by using HEV IgM and HEV IgG lab test information. HEV usually causes an acute and self-limiting infection but, can occasionally cause severe disease and/or chronic infection; our data collection approach sets out to include both categories. Patients included in the extract meet at least one of the following criteria:

a. patients with anti-HEV IgM and anti-HEV IgG both positive;
b. patients with HEV RNA detected in serum and/or stool by RT-PCR.

There are 14 patients with acute HEV infection in our current database. One of these patients is coinfected with HBV, and this patient is also diagnosed with chronic kidney disease. Among them, 4 patients are male, and gender information was missing for the other 10. The median age at the time of patients first positive HEV test was 61.5 years (range: 21-86).

## DISCUSSION

This paper describes the methodology for the NIHR HIC informatics infrastructure (pipeline) development for collating data across multiple sites, and presents initial cohorts created as part of the NIHR HIC Viral Hepatitis theme. This demonstrates that routinely collected patient data can be aggregated across multiple centres to create datasets for research. Data collected for this collaboration from one site have already been used in an analysis of HBsAg loss [9] and an analysis of data from all sites is currently underway. Further internal analysis is planned and collaborations with industry partners have been established to address specific translational research questions.

Whilst the NIHR HIC viral hepatitis theme has been able to demonstrate that routinely collected patient data can be aggregated across multiple centres to create datasets for research, challenges still remain. For example, data submission completeness differs across sites, as the original data is stored in differently and may be therefore easier to process at each site. Large amounts of imaging report and biopsy report data remains embedded in free-text which may contain patient identifiers, meaning it cannot be transmitted to the central site for processing, and each individual site has to develop free-text anonymisation protocols or perform manual extraction of this data. With these challenges in mind, optimization of the data model will be continued, and a comprehensive data dictionary continues to be updated accordingly for researchers across the participating centres and external collaborators. In addition, natural language processing (NLP) algorithms for automatically extracting data and information from free-text examination reports and clinical notes will be embedded into the data collating process, to eliminate the requirement for manual extraction.

Through the new pipeline, electronic clinical data collected for the routine care of individuals with hepatitis B, C, D, and E infection has been collated over the last three years, across five NHS trusts, and re-used for viral hepatitis research. The datasets created can be requested by internal and external research groups for analysis; these requests are reviewed and approved according to the agreed governance procedures. Data collection for the NIHR HIC viral hepatitis theme is an ongoing process, and other NHS trusts that have signed up to the NIHR HIC governance framework have been invited to join the collaboration. Collection of large datasets on viral hepatitis via the NIHR HIC programme is not only a cost-effective method of data collection but allows novel analyses to be performed, giving further insight into viral hepatitis in the UK.

## Data Availability

Data is available to researchers from on request

https://hic.nihr.ac.uk/

## ACKNOWLEDGEMENTS

The authors would like to thank all the research nurses and research admin staff at the contributing sites for their help in developing this project and Theresa Noble for her work supporting the NIHR HIC.

## REFRENCES

1 NIHR: Health Informatics Collaborative. https://hic.nihr.ac.uk/ (accessed 17 Sep 2019).

2 Miotto R, Wang F, Wang S, et al. Deep learning for healthcare: Review, opportunities and challenges. Brief Bioinform 2017;19:1236–46. doi:10.1093/bib/bbx044

3 WHO | Hepatitis B. http://who.int/mediacentre/factsheets/fs204/en/ (accessed 27 Feb 2015).

4 Stanaway JD, Flaxman AAD, Naghavi M, et al. The global burden of viral hepatitis from 1990 to 2013: findings from the Global Burden of Disease Study 2013. Lancet 2016;0:988–97. doi:10.1016/S0140-6736(16)30579-7

5 Griggs D, Stafford-Smith M, Gaffney O, et al. Policy: Sustainable development goals for people and planet. Nature. 2013;495:305–7. doi:10.1038/495305a

6 World Health Organization. *Global hepatitis report, 2017*. World Health Organization 2017. doi: ISBN 978-92-4-156545-5

7 The data collection and management for this paper was performed using the OpenClinica open source software. www.OpenClinica.com

8 Nelson EK, Piehler B, Eckels J, et al. LabKey Server: An open source platform for scientific data integration, analysis and collaboration. *BMC Bioinformatics* Published Online First: 2011. doi:10.1186/1471-2105-12-71

9 Downs LO, Smith DA, Lumley SF, et al. Electronic Health Informatics Data To Describe Clearance Dynamics of Hepatitis B Surface Antigen (HBsAg) and e Antigen (HBeAg) in Chronic Hepatitis B Virus Infection. MBio Published Online First: 2019. doi:10.1128/mbio.00699-19

